# The economic gains of tissue paper’s hygiene benefits

**DOI:** 10.64898/2026.05.19.26353582

**Authors:** Andrés Cruz, Roberta Lesma, René Kim, Mark Wilcox

**Author notes:** Contributed equally (Andrés Cruz, Roberta Lesma).

## Abstract

**Background:** The choice of hand-drying method affects microbial contamination levels but its economic consequences have not been systematically quantified.

**Methods:** By applying a quantitative microbial risk assessment framework, we translated the documented contamination differential between jet air dryers and paper towels into infection risk estimates, and embedded these into an established health economic model of healthcare-associated infections in NHS hospitals and an illustrative productivity analysis for the EU workforce.

**Results:** The median estimated avoidable HCAI cost attributable to jet air dryer presence in UK NHS clinical areas was £58 million per year, representing 2.1% of total HCAI expenditure for the affected hospital population, with a 50% certainty interval of £33-84 million. Extended to the EU workforce, the same contamination differential implied a median of €1.7 billion in € annual productivity gains, due to reduced absenteeism, for a shift to use of paper towels in public restrooms.

**Conclusions:** These findings suggest that hand-drying method selection carries measurable economic implications that are not currently reflected in facility management practice. The evidence supports the prioritisation of paper towels in clinical and public settings as a cost-effective infection control measure.

## 1 Introduction

In 1847, Semmelweis demonstrated that improving handwashing practices could dramatically reduce mortality from puerperal fever. The lesson history drew from Semmelweis - and from the long delay before his findings were accepted - is that the mechanisms of infectious disease transmission often precede scientific consensus, and that waiting for certainty carries a measurable human cost.

Hand hygiene remains a cornerstone of infection prevention. Residual moisture substantially increases microbial transfer to surfaces and other individuals (Patrick et al., 1997), and incompletely dried hands may negate much of the benefit achieved by careful washing. The method used to dry hands therefore affects not only the individual’s hygiene status but the microbial quality of the shared environment.

Three principal hand-drying methods are in common use: paper towels, warm air dryers, and jet air dryers. In most settings, the choice between them is made on the basis of cost, waste management, sustainability, and user preference. These three methods differ fundamentally in their drying mechanism: paper towels remove moisture by absorption, with the friction of towelling providing additional mechanical removal of microorganisms from the skin surface; warm air dryers evaporate moisture through heated airflow; and jet air dryers physically strip water from the hands using high-velocity air. There is a growing body of evidence suggesting that this choice also affects the risk of person and environmental contamination.

Notably, the absence of a causal link between hand-drying method and clinical infection outcomes does not constitute evidence of equivalence. It reflects a structural gap: the experiment that would definitively resolve the question has considerable challenges and may be ethically impermissible. Reynolds et al. (2021) recommended quantitative microbial risk assessment (QMRA) as the appropriate next step for evaluating the health exposure potential of different drying methods.

This study aimed to respond to that gap. Drawing on QMRA methodology - which has been successfully applied to estimate infection risk from environmental surface contamination in healthcare settings (Wilson et al., 2018) - we translated the documented microbial dispersal differentials between hand-drying methods into estimated infection risk.

We addressed this question in two stages. First, we applied the QMRA framework to estimate the differential infection risk between paper towels and jet air dryers, and embedded this differential into an established health economic model of healthcare-associated infections (HCAIs) in NHS hospitals - quantifying the avoidable economic burden attributable to the presence of jet air dryers in clinical areas. Second, we extended the same infection risk differential beyond the clinical setting, in order to estimate what the documented hygiene differential could mean at the scale of the EU workforce.

## 2 Methodology

The methodology was structured in three sequential components. The first component applied a QMRA framework to estimate how hand-drying method affects the probability of infection per exposure event, drawing on empirical contamination data and an established doseresponse relationship. The second component embedded this infection risk differential into the health economic model of Guest et al. (2019), adapting it to the NHS context to estimate the HCAI burden attributable to the use of jet air dryers in clinical areas. The third component applied the infection risk differential to worker absenteeism and productivity loss resulting from public bathroom visits.

### 2.1 QMRA framework

The (QMRA) framework estimated the differential infection risk associated with paper towels and jet air dryers as hand-drying methods. The analysis comprised three steps: characterisation of post-drying hand contamination levels by drying method; estimation of the resulting infectious dose; and derivation of the probability of infection per exposure event.

#### 2.1.1 Exposure scenario

The modelled exposure scenario was a single hand-drying event by an individual whose hands retain residual microbial contamination following handwashing. This reflects the documented reality that handwashing does not achieve sterility, and instead that a proportion of individuals retain viable pathogens on their hands post-wash. The modelled outcome is the probability of infection per hand-drying event as a function of drying method chosen.

#### 2.1.2 Contamination inputs

Post-drying hand contamination levels were derived from Redway and Fawdar (2008), who measured bacterial colony-forming units (CFU) on hands before and after drying. Measurements were taken on fingerpads specifically, as these are the regions most implicated in contact transmission (Snelling et al., 2011).

The results, summarised in Table 1, show that Paper Towel 1 reduced mean fingerpad bacterial counts by 51% across all three growth media, while Paper Towel 2 achieved a 76% reduction — suggesting that towel material and absorbency may meaningfully affect drying efficacy. Jet air dryers increased mean fingerpad bacterial counts by 42%.

**Table 1.**
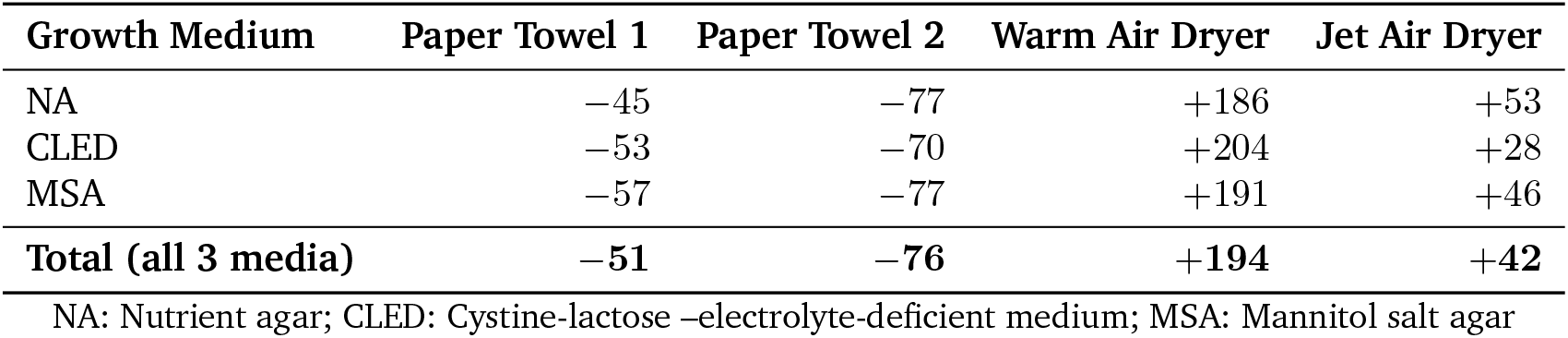
Mean percentage change (%) in bacterial counts on fingerpads before and after hand drying, by method and growth medium. Negative values indicate reductions; positive values indicate increases in bacterial numbers. Source: Redway and Fawdar (2008).

For the primary analysis, Paper Towel 1 values were used, yielding contamination factors of 0.49 for paper towels (*C*_*pt*_) and 1.42 for jet air dryers (*C*_*ad*_), applied to a postwash reference contamination level. The Paper Towel 2 results indicate that the estimate we used is probably conservative, as a more absorbent towel could produce a larger differential.

#### 2.1.3 Dose-response relationship and infection probability

We assumed a proportional relationship between hand contamination and the microbial dose available for mucosal inoculation. This assumption is supported by the near-perfect linearity observed in the Wilson et al. (2018) single-contact scenario, in which a 94.1% reduction in surface viral concentration produced a 94.1% reduction in infection risk. To translate this contamination differential into an infection risk estimate, a reference pathogen is required; i.e. a specific virus for which the relationship between dose and infection probability is well established.

Influenza A virus was used as the reference pathogen, given the availability of well-characterised dose-response parameters and its established use in surface contamination QMRA modelling (Wilson et al., 2018).

#### 2.1.4 Risk comparison

Infection probabilities were calculated for each drying method by applying the method-specific contamination factor — derived from the fingerpad bacterial count measurements described earlier — to the reference post-wash contamination level and then applying the dose-response relationship described above. Because the dose-response relationship is proportional, the contamination factors translate directly into infection risk ratios.

### 2.2 HCAI burden model

*Model structure*. The HCAI burden model we used builds on the health economic decision model published by Guest et al. (2019), which estimated the clinical and economic impact of HCAIs in an average NHS general hospital in England.

Guest et al. (2019) modelled the pathways of adult inpatients and front-line healthcare professionals (HCPs) who acquired an HCAI in a hospital setting over a period of one year.

The HCAI burden model produced several outputs: occupied hospital bed days, total annual costs, HCP absenteeism, and HCAI-related mortality. The present analysis focused on total annual costs attributable to HCAIs, reported as the most actionable measure for facility management and policy decisions. Occupied bed days were not reported directly: the heterogeneity of bed types across NHS hospitals and the variation in length of stay across HCAI types make aggregate bed day figures difficult to interpret without a level of disaggregation that the current model does not provide. HCP absenteeism and HCAI-related mortality are present in the underlying model but are similarly not reported here. Providing estimates of preventable deaths from modeling assumptions was beyond the intention of this paper and we do not consider it appropriate to do so without a level of epidemiological precision that the current model does not claim to offer.

The cost of inaction was estimated by running the Guest et al. (2019) model using the HCAI probability associated with jet air dryer presence, and paper towels only. The difference in infection cases and economic costs between these two runs represents the avoidable burden - the cost of inaction - attributable to the presence of jet air dryers in NHS clinical areas.

To derive the two HCAI probabilities, the model disaggregated the blended population probability of patients acquiring a HCAI in clinical areas, *P*_*B*_, into paper-towel-only and air-dryer-present components. The paper-towel-only and air-dryer-present probabilities of acquiting a HCAI, *P*_*pt*_ and *P*_*ad*_ respectively, and the difference between them, ∆*P*, were derived using the following equations:

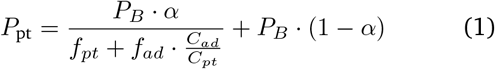

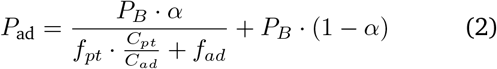

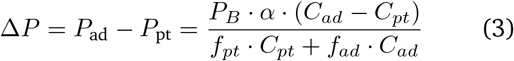

where the variables are defined as follows:

*α* Proportion of HCAI incidence attributable to hand hygiene practices

*f*_*pt*_ Proportion of NHS hospitals with paper towels only in clinical areas

*f*_*ad*_ Proportion of NHS hospitals with air dryers present in clinical areas

*C*_*pt*_ Contamination factor for paper towels

*C*_*ad*_ Contamination factor for jet air dryers

The total number of HCAIs prevented (∆*I*) by replacing jet air dryers with paper towels in all hospitals is then obtained by scaling the probability differential by the total patient population in air-dryer-present hospitals:

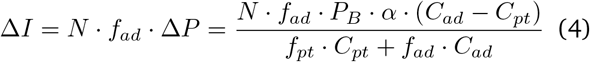

where *N* is the number of inpatients.

The prevented HCAIs is thus driven entirely by the contamination differential (*C*_*ad*_ − *C*_*pt*_), scaled by the size of the affected population *N* · *f*_*ad*_, the baseline HCAI probability *P*_*B*_, and the proportion of HCAIs attributable to hand hygiene *α*. With all fixed parameters substituted in, the expression reduced to a function of two unknowns only: *P*_*B*_ and *α*.

To characterise the full distribution of plausible outcomes, *P*_*B*_ and *α* were varied independently and uniformly across *P*_*B*_ ∈ [4.7%, 7.6%] and *α* ∈ [1%, 15%] respectively, with all other parameters held fixed at the values reported in Table 2. Since ∆*I* is a product of two independent uniform random variables scaled by a constant, its probability density can be derived exactly in closed form.

**Table 2.** Model parameters and sensitivity analysis ranges.

| Parameter | Base case | Range | Source |
| --- | --- | --- | --- |
| *HCAI probabilities* |  |  |  |
| Blended probability of patients acquiring a HCAI ($P_B$) | | 4.7%–7.6% | Guest et al. (2019); UK Health Security Agency (2023) |
| Incidence rate attributable to hand hygiene ($\alpha$) | | 1%–15% | Expert opinion |
| *Hospitals distribution* |  |  |  |
| Proportion of hospitals with paper towels only ($f_{pt}$) | 82% | Fixed | Survey of 161 NHS hospitals |
| Proportion of hospitals with air dryers present ($f_{ad}$) | 18% | Fixed | Survey of 161 NHS hospitals |
| *Contamination factors* |  |  |  |
| Paper towel factor ($C_{pt}$) | 0.49 | Fixed | Redway & Fawdar (2008) |
| Jet air dryer factor ($C_{ad}$) | 1.42 | Fixed | Redway & Fawdar (2008) |
| *Hospital parameters* |  |  |  |
| Annual adult admissions per hospital | 71,424 | Fixed | NHS Digital |
| Adult inpatient beds per hospital | 498 | Fixed | NHS Digital |
| Front-line HCPs per hospital | 5,110 | Fixed | NHS Digital |
| Total NHS annual admissions ($N$) | 14,642,000 | Fixed | NHS Digital |
| *Care pathway parameters* |  |  |  |
| Proportion of HCAI patients without extended stay ($\eta$) | 5% | Fixed | Guest et al. (2020) |
| Probability of remaining on general ward ($\pi_w$) | 90% | Fixed | Guest et al. (2019) |
| Probability of isolation following HCAI ($\pi_{iso}$) | 9.96% | Fixed | Guest et al. (2019) |
| Probability of ICU transfer following HCAI ($\pi_{icu}$) | 0.04% | Fixed | Guest et al. (2019) |
| Average increased length of stay, ward and isolation ($L_w$) | 9.1 days | Fixed | Guest et al. (2019) |
| Average additional ICU stay ($L_{icu}$) | 3.97 days | Fixed | Guest et al. (2019) |
| Additional HCP costs ($H$) | £3,784,018 | Fixed | Model output |
| *Resource costs* |  |  |  |
| General ward cost per excess bed day ($c_w$) | £351.00 | Fixed | Guest et al. (2019) |
| ICU cost per day ($c_{icu}$) | £1,621.16 | Fixed | Guest et al. (2019) |
| Isolation ward cost per day ($c_{iso}$) | £586.00 | Fixed | Guest et al. (2019) |
| NHS bank staff per day | £285.19 | Fixed | Guest et al. (2019) |
| Agency staff per day | £443.59 | Fixed | Guest et al. (2019) |
| Average cost per HCAI patient with extended stay ($\bar{c}$) | £3,414 | Fixed | Guest et al. (2019), using above-stated parameter values |

Not every patient who acquires an HCAI will experience an extended hospital stay: a proportion (*η*) will recover within their originally expected admission length and therefore generate no additional costs. Therefore, estimated avoidable cost savings (∆*C*) were obtained by applying the average cost per patient with HCAI,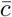, to the subset of cases prevented that are expected to result in an increased length of stay, 1 *− η*:

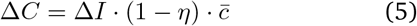

#### 2.2.1 Model parametrisation

The model was populated with the parameter values summarised in Table 2 in the Appendix. Parameters were drawn from published literature, national NHS data, and primary survey data collected for this study. Where there was uncertainty around a parameter estimate, a range was applied. All other parameters were treated as fixed. Each parameter is set out in turn below.

The baseline probability of patients acquiring a HCAI (*P*_*B*_) and the incidence rate attributable to hand hygiene (*α*) were the two parameters subject to sensitivity analysis. *P*_*B*_ ranges from 4.7% — taken from Guest et al. (2019), who derived this figure from European Centre for Disease Prevention and Control surveillance data — to 7.6% [UK Health Security Agency, 2023]. *α* ranged from 1% to 15%, reflecting the broad uncertainty around the precise contribution of hand hygiene practices to HCAI transmission.

The distribution of NHS hospitals by hand-drying method in clinical areas was derived from a survey of 161 NHS hospitals conducted as part of this study. All 161 hospitals reported the use of paper towels in clinical areas; 29 additionally reported the presence of jet air dryers in the same clinical areas, yielding *f*_*pt*_ = 0.82 and *f*_*ad*_ = 0.18. These values were treated as fixed, as the survey represents the most complete available data on hand-drying practice in NHS clinical areas in England.

The contamination factors were derived from the fingerpad bacterial count measurements reported by Redway and Fawdar (2008). Paper towels reduced mean fingerpad bacterial counts by 51% relative to post-wash levels, yielding *C*_*pt*_ = 0.49. Jet air dryers increase mean fingerpad bacterial counts by 42%, yielding *C*_*ad*_ = 1.42 (see Section 2.1.2). Both values were treated as fixed.

Hospital-level parameters - annual adult admissions, adult inpatient beds, and front-line HCPs - were updated from Guest et al. (2019) & Guest et al. (2020) to reflect current NHS reference data from NHS Digital. Total NHS annual admissions were estimated at 14,642,000, from which the patient populations in paper-towel-only and air-dryer-present hospitals were derived by applying the 82%/18% distribution and assume a constant average hospitals capacity.

The average cost per HCAI patient with extended hospital stay 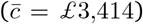was estimated from the Guest et al. (2019) model using the parameter values reported in Table 2.^1^

Resource costs followed Guest et al. (2019) with minor updates to current NHS reference costs. All cost parameters were treated as fixed. Full details are provided in Table 2 in the Appendix.

### 2.3 Avoidance of absenteeism and economic output loss

A second application of the logic presented is the estimation of avoided annual absenteeism and loss of economic output that would result from hypothetically replacing jet air dryers with paper towels in EU public restrooms. The methodology follows the same logic as the HCAI burden model: a contamination differential between drying methods was translated into an infection differential, which was then converted into an economic cost.

#### 2.3.1 Exposure and infection model

The total number of annual visits, *V*, to public restrooms by the EU workforce was estimated as:

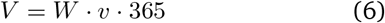

where *W* denotes the total EU workforce and *v* the average number of daily visits to restrooms per worker. Visits were distributed between air-dryer-equipped and paper-towel-equipped restrooms according to shares *γ*_*ad*_ and 1 *− γ*_*ad*_ respectively, such that *V*_*ad*_ = *γ*_*ad*_ *· V* and *V*_*pt*_ = (1 *− γ*_*ad*_) *· V*.

For each visit, the probability of developing a symptomatic infection was obtained by applying the drying-method contamination factor to a baseline post-handwashing residual contamination probability *p*_0_, and then multiplying by the probability of developing a symptomatic infection conditional on contamination *ϕ*:

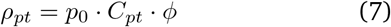

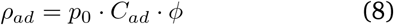

The total number of infections prevented annually, ∆*I*_*EU*_, by switching all air-dryer-equipped restrooms to paper towels is then:

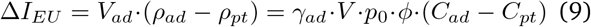

Each prevented infection was assumed to prevent *d* days of sick leave, generating a total of ∆*I*_*EU*_*· d* lost working days. The economic value *Y* of these prevented absences was estimated as:

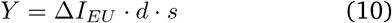

where *s* denotes the average uncompensated output loss per sick day, representing only the productive output lost and not subsequently recovered.

*d* Average number of sick leave days per infection

*s* Average uncompensated output loss per sick day

#### 2.3.2 Model parametrisation

The two parameters subject to uncertainty are *p*_0_ and *ϕ*. Both were varied independently and uniformly within their respective ranges: *p*_0_ ∈ [0.2%, 1%]^2^ and *ϕ* ∈ [2.5%, 20%]. We assumed *γ*_*ad*_ = 0.5, meaning that visits are distributed evenly between air-dryer-equipped and paper-towel-equipped restrooms. This is an assumption made for illustrative purposes, as comprehensive data on the distribution of hand-drying methods across EU public facilities does not exist and the true share may differ substantially across member states, settings, and facility types. The full parameter values are reported in Table 3 in the Appendix.

## 3 Results

### 3.1 QMRA framework

The contamination factors derived from Redway and Fawdar (2008) implied that jet air dryers left hands substantially more contaminated than paper towels following a hand-drying event. Because the dose-response relationship was assumed to be proportional, these contamination factors translated directly into infection risk ratios. The relative difference between the two contamination factors (1.42 −0.49)*/*1.42 *≈* 0.66 implied that switching from jet air dryers to paper towels was associated with a 66% reduction in infection risk per hand-drying event. This differential was independent of the absolute level of post-wash contamination and held across the full range of baseline assumptions, making it a robust input into the downstream models.

### 3.2 HCAI burden model

Using the uniform distributions of *P*_*B*_ and *α* as indicated in Section 2.2, the probability density function of estimated avoidable HCAIs is presented in Figure 1.

**Figure 1.**
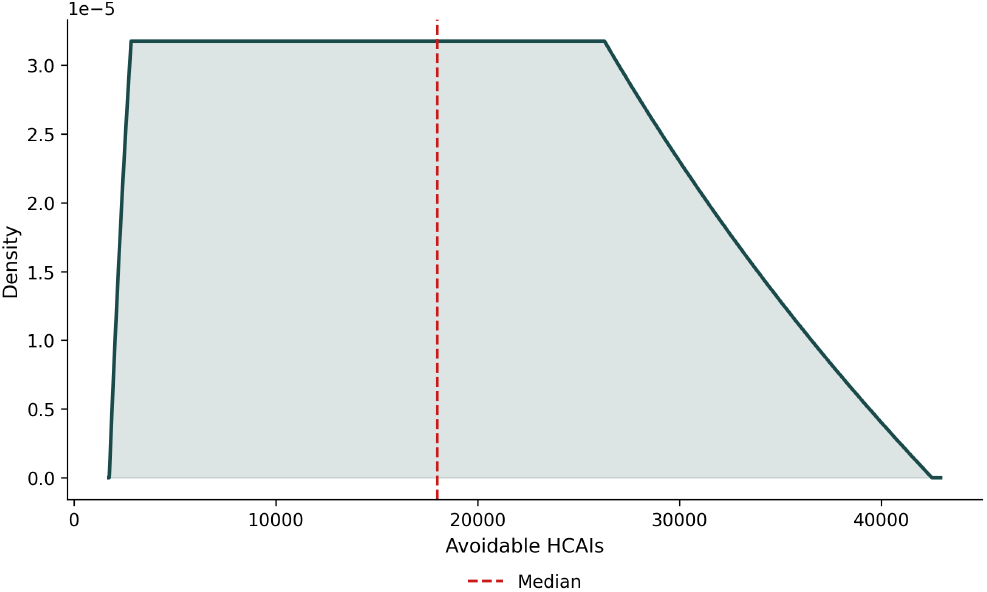
Analytical probability density of estimated avoidable HCAIs under parameter uncertainty. The distribution was derived in closed form from the exact distribution of the product of two independent uniform random variables, with *P*_*B*_ varying uniformly across *P*_*B*_ ∈ [4.7%, 7.6%] and *α* varying uniformly across *α* ∈ [1%, 15%], with all other parameters held fixed. The dashed vertical line indicates the median outcome of 18,000 avoidable HCAIs.

The probability density increased rapidly from a lower bound and plateaued across the mid-range before tapering logarithmically toward the upper bound, reflecting the characteristic shape of a product of two independent uniform random variables. The median of the distribution corresponded to 18,000 avoidable HCAIs per year. Applying 1*− η* = 0.95, this corresponded to approximately 17,100 cases expected to result in an increased length of hospital stay. At the model’s average cost of £3,414 per HCAI patient with extended stay, the median estimated avoidable cost was £58.4 million per year, representing 2.14% of total HCAI expenditure for the affected hospital population. 50% of the probability density resides between 10,100 and 25,900 avoidable HCAIs, corresponding to estimated avoidable costs of £32.8 million to £83.9 million. There is thus a 25% probability that costs are either lower or higher than this range.

### 3.3 Avoidance of absenteeism and economic output loss

Using the uniform distributions of *p*_0_ and *ϕ* as indicated in Section 2.3.2, the full distribution of estimated annual infections prevented (∆*I*_*EU*_) is presented in Figure 2.

**Figure 2.**
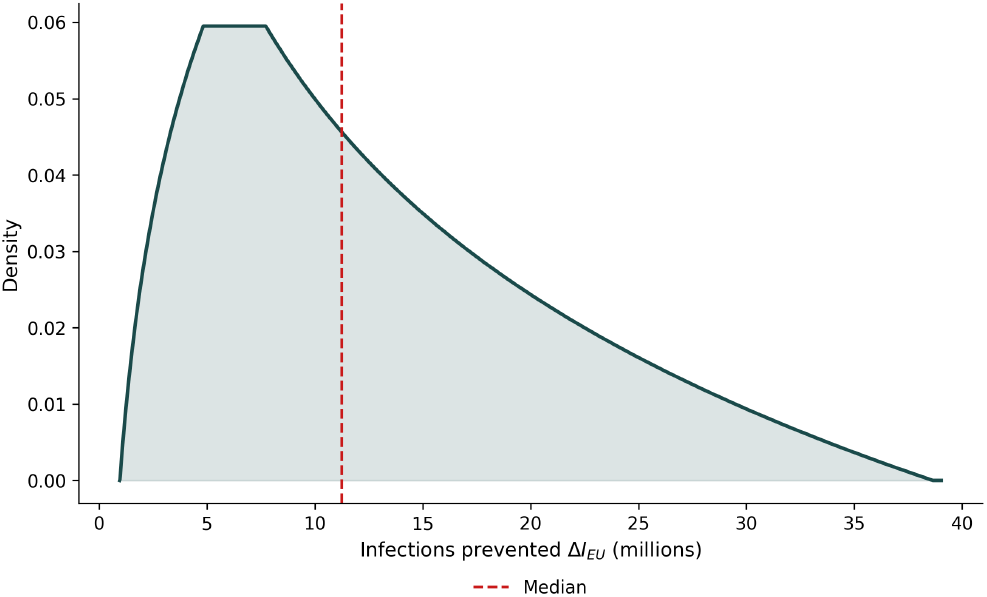
Analytical probability density of estimated annual infections prevented (∆*I*_*EU*_) under parameter uncertainty, derived from equation (9). The distribution was derived in closed form from the exact distribution of the product of two independent uniform random variables, with *p*_0_ varying uniformly across *p*_0_ ∈ [0.2%, 1%] and *ϕ* varying uniformly across *ϕ* ∈ [2.5%, 20%], with all other parameters held fixed at the values reported in Table 3. The dashed vertical line indicates the median outcome of 11.2 million infections prevented.

The distribution shares the same characteristic shape as the HCAI distribution, reflecting the identical mathematical structure of the two models. The median of the distribution corresponds to 11.2 million infections prevented annually. By assuming 3 sick-leave days per infection and an uncompensated output loss of €50 per day, this corresponds to a prevented loss of economic output of €1.69 billion. At the 50% certainty level, the estimated annual infections prevented range from approximately 6.6 million to 18.2 million, corresponding to a prevented loss of economic output of €0.99 billion to €2.73 billion respectively. These figures should be understood as illustrative rather than predictive: they demonstrate that the logical chain from hand-drying method to economic output is tractable.

**Table 3.** Economic output loss analysis parameters.

| Parameter | Base case | Range | Source |
| --- | --- | --- | --- |
| *Workforce and exposure* |  |  |  |
| EU workforce ($W$) | 167.4 million | Fixed | ILO |
| Daily public restroom visits per worker ($v$) | 0.68 | Fixed | OECD Time Use database |
| Share of restrooms with jet air dryers ($\gamma_{ad}$) | 50% | Fixed | Assumption |
| *Contamination factors* |  |  |  |
| Paper towel factor ($C_{pt}$) | 0.49 | Fixed | Redway & Fawdar (2008) |
| Jet air dryer factor ($C_{ad}$) | 1.42 | Fixed | Redway & Fawdar (2008) |
| *Infection parameters* |  |  |  |
| Baseline residual contamination probability ($p_0$) | | 0.2%–1% | Authors' assumption |
| Symptomatic infection probability ($\phi$) | | 2.5%–20% | Tsang et al. (2016); Jayasundara et al. (2014); Browne et al. (2016) |
| *Economic parameters* |  |  |  |
| Sick leave days per infection ($d$) | 3 days | Fixed | Blanchet Zumofen et al. (2023) |
| Uncompensated output loss per sick day ($s$) | €50 | Fixed | Pezzotti et al. (2019); Blanchet Zumofen et al. (2023); Brouwer et al. (2022); Hanly et al. (2020) |

*Note on symptomatic infection probability ( $\phi$ ):* There are few data on the risk of infection following a single exposure to a virus—such data invariably are obtained in laboratory or controlled conditions (Zhou et al., 2023). Multiple factors can affect infection risk, including (but not limited to) the burden of viable virus, the susceptibility and age of the exposed person, the type of virus, the route of acquisition (air, surfaces, body fluids, or a combination), and environmental humidity and temperature. The three sources cited above do not represent the same scenario as a single washroom exposure to a virus, but they provide relevant estimates of infection risk for influenza that are within or overlap with the range used here (2.5–20%).
Household secondary attack rates for PCR-confirmed influenza ranged from 1% to 38% (influenza-like illness: 6–35%; acute respiratory infection: 3–31%) (Tsang et al., 2016). A meta-regression of 34 randomised controlled trials found attack rates in adults of 3.50% (95% CI: 2.30–4.60%) for all influenza types combined, and 15.20% (95% CI: 11.40–18.90%) in children (Jayasundara et al., 2014). Influenza outbreaks aboard cruise ships affect 2–7% of passengers (Browne et al., 2016).

## 4 Discussion

The contamination differential between jet air dryers and paper towels has been documented consistently across more than a decade of independent experimental research. Redway and Fawdar (2008) established the fingerpad bacterial count differentials that underpin the present analysis. Best et al. (2014) subsequently demonstrated that airborne bacterial counts in the immediate vicinity of jet air dryer use were 27-fold higher than during paper towel drying, with bacteria persisting in the air for at least 15 minutes. Kimmitt and Redway (2016) found that jet air dryers dispersed over 1,300-fold more viral particles than paper towels at distances of up to 3 metres. Best et al. (2018), in the largest multicentre study of its type conducted across hospitals in the UK, France and Italy, documented significantly higher recovery of faecal-associated and antibiotic-resistant bacteria, including MRSA and ESBL-producing organisms, from dryer surfaces and floors in washroom using jet air dyers compared with those using paper towels. Most recently, Moura et al. (2025) extended this evidence to newer electric hand dryer models, finding that wall contamination was up to 78 times higher with high-speed jet air dryers compared to paper towels, and that aerosolised particles remained detectable on facemasks at one metre distance for up to 30 minutes following hand drying. The use of facemasks in this study, and the demonstration of contamination thereon, acted as a proxy for virus inhalation risk. The directional consistency of this evidence base, across different research groups, microbial models, and experimental designs, is striking. What was hitherto missing is a principled method for converting it into the language that facility managers and policymakers use: costs, productivity, and economic burden. The present study aimed to fill this gap.

At the median of a conservative analytical distribution, the estimated avoidable HCAI costs attributable to the presence of jet air dryers in NHS clinical areas amounted to £58.4 million per year, representing 2.14% of total HCAI expenditure for the affected hospital population.

These figures are grounded in the same experimental record that has accumulated since the WHO recommended paper towels as the preferred hand-drying method in healthcare settings in 2009. Sixteen years later, 18% of NHS hospitals still have jet air dryers present in clinical areas. The present analysis suggests that this inertia carries a quantifiable price. The 50% certainty interval for the HCAI result spans £32.8 million to £83.9 million, and under no parameter combination did the model produce a result in which air dryers were associated with lower costs than paper towels.

Beyond clinical settings, at the scale of the entire EU workforce, the same contamination differential could well prevent the loss of €1.69 billion of economic output due to absenteeism. Sick leave across EU member states has been rising steadily for over a decade. Germany offers a stark illustration: the average number of sick leave days per employee climbed from 9.5 in 2013 to 15.1 in 2023 (Statistisches Bundesamt, 2025). Although illness is due to many reasons, hand hygiene remains one of the most consistently identified drivers of infectious disease transmission in both healthcare and community settings. The hand-drying choice made in a public restroom is a small but non-trivial part of that picture, and the productivity analysis presented here suggests that its aggregate economic footprint is measurable. The evidence reviewed in this study suggests that the choice between paper towels and jet air dryers is not neutral from an infection risk perspective, with resulting economica impacts.

Several limitations of the present analysis must be acknowledged. First, the QMRA framework relies on an extrapolation of laboratory-measured contamination differentials to real-world infection outcomes. A purpose-built QMRA model for hand-drying in healthcare settings would in principle offer greater precision, but would require modelling a large number of additional variables, including aerosol dispersion dynamics, surface persistence, and contact transmission pathways, each carrying its own uncertainty. The complexity of such a model would be considerable, and the marginal gain over an approach grounded in directly comparable existing studies is not self-evident. The present analysis follows the methodology recommended by Reynolds et al. (2021) and builds on work that has been directly applied in analogous healthcare settings, representing a principled and tractable approximation in the absence of a purpose-built model.

Second, the incidence rate *α*, representing the proportion of HCAI risk attributable to hand hygiene practices, was based on expert opinion and carries the greatest uncertainty in the HCAI model. Although a sensitivity range of 1% to 15% was applied to reflect this uncertainty, the true value of this parameter remains difficult to isolate empirically from other infection control factors.

Third, the assumption that 50% of EU public restrooms are equipped with jet air dryers is unverified at the European level. Comprehensive data on the distribution of hand-drying methods across EU public facilities does not exist, and the true share may differ substantially across member states, settings, and facility types.

In addition the choice of a precise value for the uncompensated output loss per sick day (*s*) is inherently difficult to defend. European estimates in the literature range from approximately €18 to €1109 per day depending on the methodological approach and population studied. In the absence of a single authoritative estimate, we adopt a somewhat conservative value of €150. To the extent that the true value of *s* is higher, the economic output estimates reported here understate the true economic benefit of switching to paper towels.

Finally, both analyses exclude second-order effects. Both the HCAI and absenteeism models capture the direct effect on infections but do not cover infection from subsequent transmission and the associated healthcare system costs and productivity losses. To the extent that these effects are positive, the estimates reported here understate the true value of switching to paper towels.

## In conclusion

The results of our study are grounded in a simple observation: the contamination differential between jet air dryers and paper towels, documented consistently in multiple independent studies, has quantifiable economic consequences that current facility management practice does not account for. Embedded in an established health economic model of HCAI burden, this differential translated into an estimated £58.4 million in avoidable costs per year for NHS hospitals with jet air dryers present in clinical areas, representing 2.14% of total HCAI expenditure for the affected population. Extended to the EU workforce, the same differential implied €1.69 billion in annual productivity gains from a shift toward paper towels in public restrooms. While these figures are estimates rather than precise measurements, the direction of the finding was unambiguous across the full parameter space: under no combination of plausible assumptions did the model produce a result in which jet air dryers were associated with lower costs than paper towels.

The methodological contribution of this paper is the translation of a well-documented microbiological differential into a quantitative economic framework, using analytical distributions derived from probabilistic reasoning. The contamination data on which these estimates rest were not generated by this study. They reflect a body of experimental evidence accumulated over more than a decade by independent research groups. What this paper adds is a transparent, reproducible, and conservative method for converting that evidence into actionable economic terms.

The evidence presented here suggests that procurement decisions in NHS hospitals and EU public facilities could be usefully informed by these findings. Notably, replacing jet air dryers with paper towels in clinical areas does not require new regulation, clinical protocols, or capital investment.

## Competing interests

This study was funded by the European Tissue Symposium (ETS). Andrés Cruz, Roberta Lesma, and René Kim are employees of Steward Redqueen, which was commissioned by ETS to conduct this study. Mark Wilcox is Professor of Medical Microbiology at the University of Leeds and Consultant Microbiologist at Leeds Teaching Hospitals NHS Trust. Mark Wilcox is associated with ETS but is not a member of the organisation. All authors declare that the funder had no role in study design, data collection, analysis, interpretation of results, or the decision to submit for publication.

## Funding

This study was funded by the European Tissue Symposium (ETS). The funder commissioned the study but had no role in data collection, analysis, interpretation of results, or the decision to submit the manuscript for publication.

## Data availability

All data from published sources used in this study are publicly available via the cited references. Data from the survey of NHS hospitals conducted by Kimberly-Clark and the European Tissue Symposium are available from the corresponding author on reasonable request.

## Appendix

## Footnotes

1 Specifically, 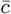 was computed as: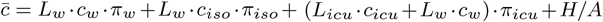where *A* = *N· P*_*B*_*·* (1*− η*) denotes the number of inpatients who acquired a HCAI and experienced an extended hospital stay. Substituting the parameter values from Table 2 yielded a ward component of £2,874.69, an isolation component of £531.13, an ICU component of £3.85, and an HCP component of £4.33, summing to £3,414.

2 The lower bound of *p*_0_=0.2% was derived from the observation that a single 3-minute bathroom visit represents approximately 0.2% of a working day (3/1440 minutes), providing a time-exposure floor for residual contamination risk. The upper bound of 1% reflects the welldocumented elevated microbial load of public bathroom environments relative to general indoor settings, assumed to be up to five times higher.

## Notes

### Competing Interest Statement

Competing interests
This study was funded by the European Tissue Sympo-
sium (ETS). Andres Cruz, Roberta Lesma, and Rene Kim are employees of Steward Redqueen, which was commissioned by ETS to conduct this study. Mark Wilcox is Professor of Medical Microbiology at the University of Leeds and Consultant Microbiologist at Leeds Teaching Hospitals NHS Trust. Mark Wilcox is associated with ETS but is not a member of the organisation. All authors declare that the funder had no role in study design, data collection, analysis, interpretation of results, or the
decision to submit for publication.

### Summary of Updates

This version of the manuscript replaces the previously uploaded file, which appeared truncated. The revised version provides the complete manuscript with all intended sections, content, and associated files included.

